# Association between having a pregnancy complicated by vaginal bleeding and risk of cancer

**DOI:** 10.1101/2021.12.29.21268235

**Authors:** Elena Dudukina, Erzsébet Horváth-Puhó, Henrik Toft Sørensen, Vera Ehrenstein

**Author notes:** **Corresponding author:** Elena Dudukina, Olof Palmes Alle 43-45, 8200, Aarhus N, Denmark.

## Abstract

**Background:** A full-term pregnancy is associated with a lower cancer risk. The risk of cancer in women with vaginal bleeding in pregnancy is unknown.

**Methods:** We conducted a registry-based cohort study (1995-2017) in Denmark. We included pregnancies (n=37 082) affected by vaginal bleeding (VB) within 20 gestational weeks among 35 514 women, VB-unaffected pregnancies (n=1 363 614) among 783 314 women, pregnancies ending in a termination (n=324 328) among 239 638 women or miscarriage (n=137 104) among 121 353 women. We computed absolute risk of cancer and hazard ratios (HR) with 95% confidence intervals (CI) adjusted for age, calendar year, morbidity, and socioeconomic factors using Cox proportional hazards regression.

**Results:** At the end of the 24-year follow-up, there were 1 320 events among VB-affected cohort, 40 420 events among VB-unaffected cohort, 10 300 events among termination cohort, and 4 790 events among miscarriage cohort. The HR for any cancer was 1.03 (95% CI: 0.97-1.08) when comparing VB-affected vs VB-unaffected pregnancies, 1.03 (95% CI: 0.97-1.09) vs terminations, and 0.90 (95% CI: 0.84-0.95) vs miscarriages. Similar results were obtained for site-specific cancers.

**Conclusions:** We found no strong evidence for an association between vaginal bleeding in pregnancy and an increased risk of cancer.

## INTRODUCTION

Gestation is sustained with progesterone, which may play a protective role for hormone-related cancers, such as endometrial and breast cancer (1,2). Threatened miscarriage and completed miscarriage share several biologic mechanisms, including increased levels of pro-inflammatory cytokines (3–5) and decreased levels of progesterone (6,7). Threatened miscarriage affects up to 30% (8–12) of clinically identified pregnancies and manifests as vaginal bleeding (VB) before 20 gestational weeks of a viable intrauterine pregnancy (8,9,13). About 30-50% (6,14,15) of VB-affected pregnancies may progress to a completed miscarriage (16–18). Previously reported risk factors for VB in pregnancy are advanced women’s age at pregnancy (≥35 years), obesity, smoking, alcohol abuse, inflammation, and low plasma levels of progesterone in gestation (16,19–26).

A full-term pregnancy is associated with a 40% reduction in risks of breast, endometrial, and ovarian cancer (27,28). Pregnancies with a duration of 33 gestational week or more were not associated with increased risk of breast cancer in contrast to pregnancies with shorter gestation (1,2,27). In line with this, a full-term pregnancy is associated with a 40% cancer risk reduction (27,28); however, reduction in cancer risk after a pregnancy ending in a miscarriage is only 10% (1). The long-term risk of cancer in women with a pregnancy complicated by VB within 20 gestational weeks is unknown and could be increased due to underlying inflammatory pathways and reduced level of progesterone. In this cohort study, we investigated an association between having a VB-affected pregnancy ending in childbirth with the subsequent risk of cancer as compared with VB-unaffected pregnancy ending in childbirth, pregnancy ending in a termination or pregnancy ending in miscarriage.

## METHODS

### Setting, design and data sources

This was a registry-based cohort study conducted using individual-level linked data from Danish health and administrative registries, set within Denmark’s universal and tax-funded health care system (29). The Civil Registration System assigns personal identifiers and contains data on date of birth, vital and civil status, and dates of immigration and emigration (29,30). Data on all deliveries are captured by the Medical Birth Registry (MBR) starting in 1973 and includes women’s and pregnancy-related characteristics including but not limited to women’s age at delivery, gestational age, and parity (31,32). Smoking status in pregnancy is collected by the MBR from 1991 and body-mass index (BMI) from 2004. Information on stillbirths at the gestational age of at least 28 full weeks is available in the MBR before April 2004 and at 22 full gestational weeks afterwards; livebirths are recorded regardless of gestational age. Information on highest education level (33), employment status (34) and personal income (35) are stored at Statistics Denmark and available starting in 1980-1981 (36). Hospital encounters are recorded in the Danish National Patient Registry (DNPR) (37,38), including data on medical procedures and discharge diagnoses, coded using the International Classification of Diseases, 8^th^ revision (ICD-8) in 1977-1993 and 10^th^ revision (ICD-10) thereafter (37). Hospital admissions are available in the DNPR from 1977, while outpatient clinic and emergency room visits are recorded from 1995 onwards (37,38). Admissions and outpatient specialist visits due to psychiatric conditions are captured as part of the the DNPR (39) and were available for this study from 1995. The Danish National Prescription Registry (40) captures data on prescribed medication dispensings from 1995 onwards and records the Anatomical Therapeutic Classification (ATC) codes of the dispensed drugs and the dates when the prescriptions were redeemed. The Danish Cancer Registry (DCR) captures all primary incident cancer diagnoses (with compulsory reporting from 1987) and contains data on tumour morphology (41).

### Study population

Using the MBR, we identified all pregnancies ending in a liveborn or stillborn delivery between 1995 and 2017. We kept one record per delivery of a multifetal pregnancy. Using the DNPR, we obtained and linked data on VB within 20 gestational weeks starting from the last menstrual period date, computed by subtracting gestational age at delivery from the delivery date (Table 1 in the electronic supplementary material). Pregnancies ending in a delivery without a VB record in the DNPR within 20 weeks of gestation were classified as VB-unaffected. The date of delivery served as the start of the follow-up for both VB-affected and VB-unaffected cohorts of pregnancies.

**Table 1.** Descriptive characteristics of the VB-affected pregnancy and comparator cohorts, Denmark, 1995-2017.

|  | Exposed cohort |  | Comparator cohorts |  |
| --- | --- | --- | --- | --- |
|  | VB-affected pregnancy, N = 37 082 | VB-unaffected pregnancy, N = 1 363 614 | Termination, N = 324 328 | Miscarriage, N = 137 104 |
| Women, N | 35 514 | 783 314 | 239 638 | 121 353 |
| Characteristic | n (%)<sup>a</sup> | n (%)<sup>a</sup> | n (%)<sup>a</sup> | n (%)<sup>a</sup> |
| Age at pregnancy end, years |  |  |  |  |
| <20 | 565 (1.5) | 20 010 (1.5) | 46 215 (14.0) | 4 635 (3.4) |
| 20-24 | 5 030 (14.0) | 163 880 (12.0) | 74 390 (23.0) | 17 410 (13.0) |
| 25-29 | 12 265 (33.0) | 464 200 (34.0) | 68 090 (21.0) | 37 455 (27.0) |
| 30-34 | 12 380 (33.0) | 476 500 (35.0) | 65 425 (20.0) | 39 820 (29.0) |
| 35-39 | 5 720 (15.0) | 202 705 (15.0) | 49 490 (15.0) | 26 205 (19.0) |
| 40+ | 1 125 (3.0) | 36 320 (2.7) | 20 725 (6.4) | 11 575 (8.4) |
| Calendar year of pregnancy end |  |  |  |  |
| 1995-1999 | 12 660 (34.0) | 308 315 (23.0) | 88 940 (27.0) | 33 565 (24.0) |
| 2000-2004 | 8 265 (22.0) | 243 075 (18.0) | 63 420 (20.0) | 25 470 (19.0) |
| 2005-2008 | 7 220 (19.0) | 304 450 (22.0) | 72 305 (22.0) | 33 885 (25.0) |
| 2009-2013 | 4 730 (13.0) | 229 010 (17.0) | 50 600 (16.0) | 25 365 (19.0) |
| 2013-2017 | 4 205 (11.0) | 278 765 (20.0) | 49 070 (15.0) | 18 820 (14.0) |
| Smoking in pregnancy |  |  |  |  |
| Smoker | 6 990 (19.0) | 209 855 (15.0) | Not available |  |
| Non-smoker | 26 205 (71.0) | 1 049 620 (77.0) |  |  |
| Missing | 6 990 (19.0) | 209 855 (15.0) |  |  |
| Parity at the index date<sup>b</sup> |  |  |  |  |
| Nulliparous | 0 (0.0) | 0 (0.0) | 205 310 (63.0) | 110 460 (81.0) |
| Primiparous | 17 105 (46.0) | 629 085 (46.0) | 32 775 (10.0) | 10 890 (7.9) |
| Multiparous | 19 980 (54.0) | 734 530 (54.0) | 86 245 (27.0) | 15 750 (11.0) |
| Pregnancy number at the index date |  |  |  |  |
| 1 | 11 300 (30.0) | 512 700 (38.0) | 112 340 (35.0) | 43 995 (32.0) |
| 2 | 11 545 (31.0) | 447 730 (33.0) | 59 810 (18.0) | 39 945 (29.0) |
| 3 | 7 045 (19.0) | 232 420 (17.0) | 58 840 (18.0) | 25 525 (19.0) |
| 3+ | 7 190 (19.0) | 170 765 (13.0) | 93 340 (29.0) | 27 640 (20.0) |
| Civil status |  |  |  |  |
| Married or in partnership | 22 330 (60.0) | 833 375 (61.0) | 102 500 (32.0) | 68 615 (50.0) |
| Not married | 10 530 (28.0) | 387 465 (28.0) | 165 230 (51.0) | 43 105 (31.0) |
| Missing | 4 225 (11.0) | 142 775 (10.0) | 56 600 (17.0) | 25 385 (19.0) |
| Employment status |  |  |  |  |
| Employed | 26 970 (73.0) | 1 017 935 (75.0) | 194 860 (60.0) | 98 695 (72.0) |
| Retirement or state pension | 3 395 (9.2) | 100 590 (7.4) | 41 220 (13.0) | 13 030 (9.5) |
| Unemployed | 5 750 (16.0) | 201 410 (15.0) | 80 905 (25.0) | 21 330 (16.0) |
| Missing | 965 (2.6) | 43 680 (3.2) | 7 340 (2.3) | 4 055 (3.0) |
| Highest achieved education |  |  |  |  |
| Basic education | 9 815 (26.0) | 272 030 (20.0) | 145 845 (45.0) | 35 680 (26.0) |
| High school or similar | 15 095 (41.0) | 553 485 (41.0) | 109 340 (34.0) | 53 405 (39.0) |
| Higher education (bachelor's degree, bachelor's degree equivalent or higher) | 10 605 (29.0) | 473 525 (35.0) | 48 940 (15.0) | 41 135 (30.0) |
| Missing | 1 570 (4.2) | 64 575 (4.7) | 20 205 (6.2) | 6 885 (5.0) |
| Year-specific income, quartiles |  |  |  |  |
| Q1 | 8 895 (24.0) | 282 190 (21.0) | 130 535 (40.0) | 33 130 (24.0) |
| Q2 | 9 460 (26.0) | 326 595 (24.0) | 82 285 (25.0) | 34 490 (25.0) |
| Q3 | 8 870 (24.0) | 359 000 (26.0) | 52 580 (16.0) | 32 695 (24.0) |
|  | VB-affected pregnancy, N = 37 082 | VB-unaffected pregnancy, N = 1 363 614 | Termination, N = 324 328 | Miscarriage, N = 137 104 |
| Women, N | 35 514 | 783 314 | 239 638 | 121 353 |
| Characteristic | n (%)<sup>a</sup> | n (%)<sup>a</sup> | n (%)<sup>a</sup> | n (%)<sup>a</sup> |
| Q4 | 9 185 (25.0) | 362 210 (27.0) | 47 165 (15.0) | 33 220 (24.0) |
| Missing | 670 (1.8) | 33 620 (2.5) | 11 760 (3.6) | 3 565 (2.6) |
| **Origin** |  |  |  |  |
| Danish | 30 930 (83.0) | 1 158 025 (85.0) | 273 430 (84.0) | 114 750 (84.0) |
| Non-Danish | 6 145 (17.0) | 205 150 (15.0) | 50 900 (16.0) | 22 355 (16.0) |
| Missing | 5 (<0.1) | 445 (<0.1) | - | - |
| **Reproductive factors history** |  |  |  |  |
| At least one VB diagnosis | 4 870 (13.0) | 66 355 (4.9) | 23 940 (7.4) | 17 005 (12.0) |
| At least one VB diagnosis within 20 gestational weeks of a pregnancy ending in a delivery | 2 320 (6.3) | 29 305 (2.1) | 12 000 (3.7) | 5 040 (3.7) |
| At least one termination | 8 610 (23.0) | 224 450 (16.0) | 110 480 (34.0) | 30 195 (22.0) |
| At least one miscarriage | 8 120 (22.0) | 165 775 (12.0) | 38 645 (12.0) | 20 940 (15.0) |
| Hypertensive disorders of pregnancy | 1 345 (3.6) | 45 100 (3.3) | 10 920 (3.4) | 5 525 (4.0) |
| Placenta praevia | 95 (0.3) | 3 480 (0.3) | 1 040 (0.3) | 475 (0.3) |
| Abruptio placentae | 255 (0.7) | 5 995 (0.4) | 2 090 (0.6) | 930 (0.7) |
| Gestational diabetes | 280 (0.8) | 11 020 (0.8) | 2 570 (0.8) | 1 550 (1.1) |
| **Reproductive factors in a current pregnancy** |  |  |  |  |
| Hypertensive disorders of pregnancy | 1 610 (4.3) | 51 415 (3.8) | Not available |  |
| Eclampsia superimposed on preexisting hypertension | 10 (<0.1) | 305 (<0.1) | Not available |  |
| Eclampsia | 20 (<0.1) | 610 (<0.1) | Not available |  |
| Preeclampsia | 1 110 (3.0) | 34 730 (2.5) | Not available |  |
| Gestational hypertension | 485 (1.3) | 18 490 (1.4) | Not available |  |
| Gestational proteinuria | 320 (0.9) | 8 345 (0.6) | Not available |  |
| Placenta praevia | 475 (1.3) | 7 445 (0.5) | Not available |  |
| Abruptio placentae | 385 (1.0) | 7 110 (0.5) | Not available |  |
| Gestational diabetes | 790 (2.1) | 25 375 (1.9) | Not available |  |
| Gestation age of a new-born, weeks (median, 25<sup>th</sup>-75<sup>th</sup> percentile) | 39.0 (38.0, 40.0) | 40.0 (39.0, 41.0) | Not available |  |
| **Comorbidities** |  |  |  |  |
| Obesity diagnosis | 2 480 (6.7) | 103 560 (7.6) | 11 355 (3.5) | 6 135 (4.5) |
| PCOS | 405 (1.1) | 12 215 (0.9) | 1 150 (0.4) | 1 185 (0.9) |
| Thyroid disorders | 915 (2.5) | 30 135 (2.2) | 5 065 (1.6) | 2 945 (2.1) |
| COPD | 450 (1.2) | 14 140 (1.0) | 3 415 (1.1) | 1 450 (1.1) |
| Chronic kidney diseases | 115 (0.3) | 2 755 (0.2) | 770 (0.2) | 305 (0.2) |
| Chronic liver diseases | 100 (0.3) | 3 330 (0.2) | 950 (0.3) | 385 (0.3) |
| Hyperlipidemia | 120 (0.3) | 4 085 (0.3) | 1 045 (0.3) | 580 (0.4) |
| Diabetes types 1 and 2 | 760 (2.0) | 26 795 (2.0) | 3 390 (1.0) | 2 775 (2.0) |
| Hypertension | 235 (0.6) | 7 575 (0.6) | 1 600 (0.5) | 915 (0.7) |
| Any connective tissue disorder | 2 930 (7.9) | 71 165 (5.2) | 18 715 (5.8) | 8 410 (6.1) |
| Any psychiatric condition identified using diagnostic codes | 5 145 (14.0) | 140 635 (10.0) | 61 995 (19.0) | 17 730 (13.0) |
| Any psychiatric condition identified using diagnostic codes and psychotropic medication proxy | 7 595 (20.0) | 225 080 (17.0) | 87 280 (27.0) | 28 275 (21.0) |
| **Medication use history** |  |  |  |  |
| NSAIDs<sup>c</sup> | 3 135 (8.5) | 87 400 (6.4) | 49 615 (15.0) | 21 670 (16.0) |
| Steroids for systemic use<sup>d</sup> | 2 845 (7.7) | 102 390 (7.5) | 20 780 (6.4) | 11 380 (8.3) |
|  | **VB-affected pregnancy,**<br>**N = 37 082** | **VB-unaffected pregnancy,**<br>**N = 1 363 614** | **Termination,**<br>**N = 324 328** | **Miscarriage,**<br>**N = 137 104** |
| **Women, N** | **35 514** | **783 314** | **239 638** | **121 353** |
| **Characteristic** | **n (%)<sup>a</sup>** | **n (%)<sup>a</sup>** | **n (%)<sup>a</sup>** | **n (%)<sup>a</sup>** |

<sup>a</sup> All counts are rounded to the nearest 5 to adhere to Denmark Statistics data confidentiality requirements; percentages $\geq 10\%$ are rounded to the nearest whole number and percentages $< 10\%$ are rounded to the first decimal place
<sup>b</sup> Parity was included in the statistical models as nulli- or primiparous vs multiparous
<sup>c</sup> At least one prescription 365 days before 12 gestational weeks for VB-affected and VB-unaffected pregnancies and before the index date for terminations and miscarriages
<sup>d</sup> At least one prescription any time before 12 gestational weeks for VB-affected and VB-unaffected pregnancies and before the index date for terminations and miscarriages
COPD, chronic obstructive pulmonary disease; NSAIDs, non-steroid anti-inflammatory disease; PCOS, polycystic ovary syndrome

Using the DNPR, we identified pregnancies ending in a miscarriage (spontaneous abortion) or a termination (medical or surgical abortion) in 1995-2017, using relevant diagnostic and procedure codes (Table 1 in the electronic supplementary material). To avoid multiple counting of the same miscarriages or terminations, a woman with repeated event-qualifying diagnostic or procedure code was allowed to re-enter the cohort after 180 days, whereby she was assumed to have a new pregnancy. When multiple dates associated with the pregnancy termination or miscarriage were available (date of procedure, date of hospital admission, date of hospital discharge), we included the earliest. The date of the hospital-based encounter for pregnany termination or miscarriage served as the index date. We excluded from the study pregnancy records of women with cancer or HIV/AIDS diagnosis before the index date, records with missing women’s age at the pregnancy end, and pregnancies ending in childbirth with missing gestational age. The flow-chart for study population construction is presented in Figure 1.

**Figure 1.**
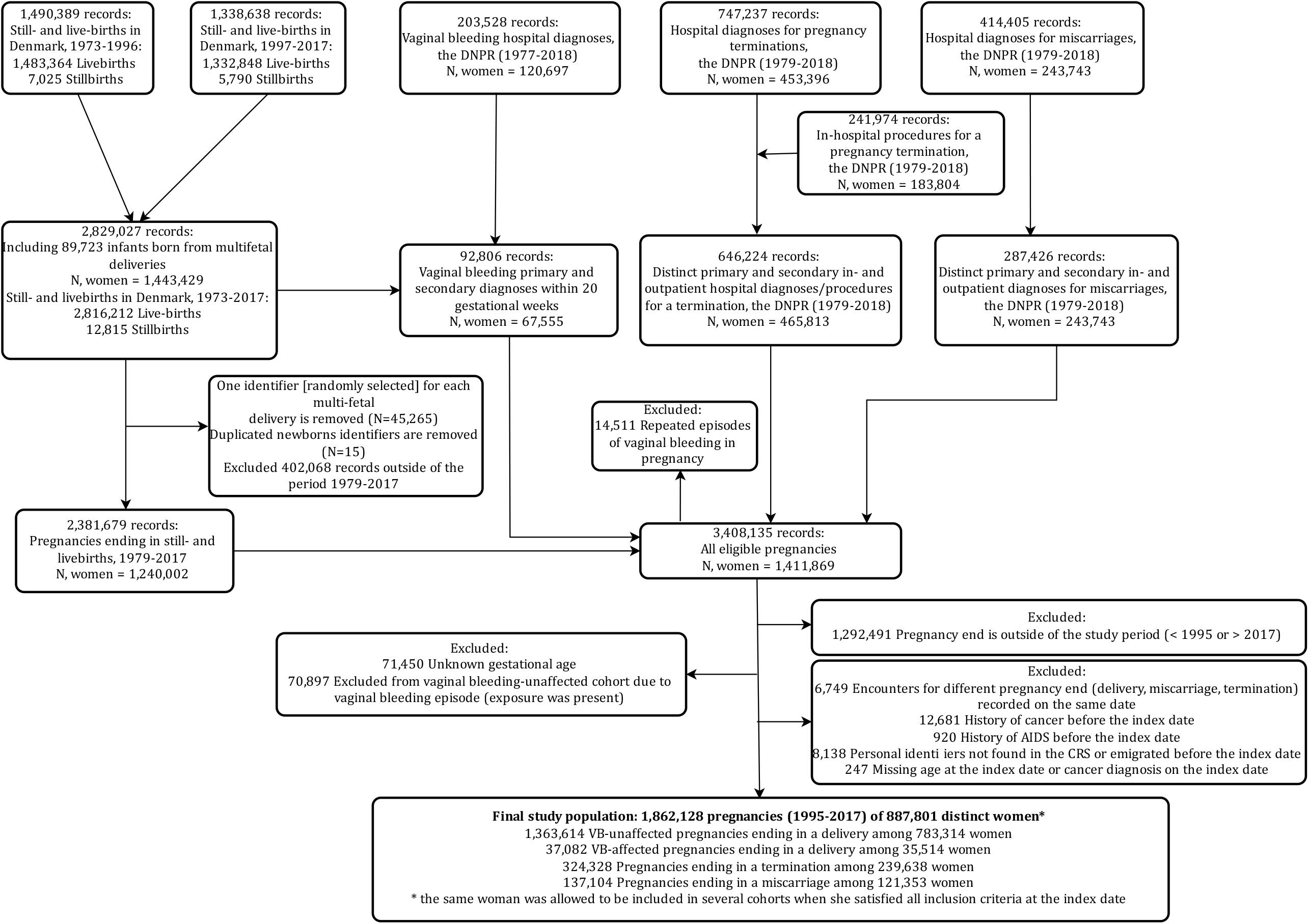
Flow chart for the study population ascertainment. Standalone figure

### Cancer outcomes

Using the DCR, we ascertained incident diagnoses of cancer. We investigated the long-term risk of any cancer; cancers at specific anatomic sites, focusing on breast, cervical, ovarian and fallopian tube, and uterine cancer, and cancers grouped as hormone-related, haematological, immune-related, smoking or alcohol-related, obesity-related, and cancers of neurological origin, as previously described (42–44)(Table 1 in the electronic supplementary material).

### Covariables

The potential confounding factors were age, calendar year of pregnancy end, women’s parity, number of previous identifiable pregnancies, socioeconomic factors (education, employment, and year-specific income level divided into quartiles), reproductive factors (pre-index date vaginal bleeding, termination, or miscarriages, placental complications in previous and current deliveries), chronic somatic conditions (obesity, polycystic ovary syndrome, thyroid disorders, chronic kidney disease, chronic liver disease, hyperlipidaemia, diabetes, hypertension, and connective tissue disorder), and history of any psychiatric condition using the entire available lookback period. We additionally adjusted for the use of NSAIDs and steroids for systemic use before the 12 weeks of gestation for VB-affected and VB-unaffected pregnancies and before the index date for terminations and miscarriages using 365 days lookback period for NSAIDs and the entire available lookback period for steroids for systemic use. In the comparisons of pregnancies ending in childbirth, we additionally controlled for placental complications (hypertensive disorders of pregnancy, placenta praevia, and abruptio placenta) not only at the previous deliveries but also at the index pregnancy as a late manifesting proxy of potential placental dysfunction early in pregnancy.

### Statistical analyses

We followed each woman from the index date until the earliest of cancer diagnosis, death, emigration, or end of the study on 31 December 2018. We constructed the cumulative incidence curves of any cancer at the end of a 24-year follow-up while treating death and emigration as competing events (Figure 1 in the electronic supplementary material) and calculated cancer incidence rates (IRs) per 10 000 person-years (PYs). We used Cox proportional-hazards regression to compute crude and adjusted hazard ratios (HRs) for the cancer outcomes. In the analyses of all pregnancies, with each subsequent pregnancy of a woman, we accounted for the reproductive history, including prior exposure status, at the previous pregnancies. We computed robust 95% confidence intervals (CIs) taking into account that different non–independent pregnancies of the same woman were analysed (45). Further, we repeated the main analyses following women’s first identifiable pregnancy. In separate sets of analyses, we compared the risk of cancer outcomes following a VB-affected pregnancy vs 1) VB-unaffected pregnancy, 2) termination, or 3) miscarriage.

The HRs were adjusted for potential confounders (women’s age at pregnancy end, calendar period, socioeconomic factors [education, employment, and year-specific income level divided into quartiles], reproductive history [parity, number of pre-index date identifiable pregnancies, history of VB, terminations or miscarriages, hypertensive disorders of pregnancy, placenta praevia, abruptio placentae] and placenta-related complications at the current pregnancy ending in delivery [preeclampsia-eclampsia, placenta praevia, abruptio placentae]; pre-index date thyroid conditions, obesity, polycystic ovary syndrome, chronic obstructive pulmonary disease, chronic liver and kidney diseases, connective tissue disorders and medication use [non-steroid anti-inflammatory drugs and steroid for systemic use]). In analyses of all identifiable pregnancies of a woman, we ascertained the history of all covariables before the index date in a time-varying manner updating the covariables’ status at each subsequent pregnancy.

In a sensitivity analysis, we investigated the association of VB-affected pregnancy with any cancer and hormone-related, haematological, immune-related, smoking or alcohol-related, obesity-related, and cancers of neurological origin additionally adjusted for women’s BMI in analyses of pregnancies ending in a delivery; however, the follow-up was short and accrued few events. We additionally investigated the associations with cancer outcomes restricting the data to women having their last recorded pregnancy at the age of 40 years or later as the age, when most women likely reach the desired family size. We did not impute missing values and included variables with missing data in the statistical models using “missing” as a category.

We used log-minus-log plots to evaluate the proportionality of hazards assumptions, and the curves were close to parallel over the follow-up period for overall cancer. All analyses were conducted on the servers of Statistics Denmark (36) using RStudio (46) and R software for statistical computing (47) versions 3.6.0-4.1.0, including the following packages and their dependencies: tidyverse (48), ggplot2 (49), survival (45), cmprsk (50), prodlim (51), and epiR (52). Reported counts were masked to protect personal data, in compliance with the Statistics Denmark regulations.

### Ethics

Patient consent or ethical approval is not required for registry-based research according to Danish legislation. This study was reported to the Danish Data Protection Agency (53) through registration at Aarhus University (record number: AU-2016-051-000001, sequential number 605).

## RESULTS

### Descriptive characteristics

There were 37 082 VB-affected pregnancies among 35 514 women; 1 363 614 VB-unaffected pregnancies among 783 314 women; 324 328 terminations among 239 638 women and 137 104 miscarriages among 121 353 women in 1995-2017 (Figure 1). Women’s age, parity, civil status, employment, income, obesity, chronic pulmonary, chronic liver and kidney conditions, alcohol abuse, and medication use had similar distributions for VB-affected and VB-unaffected pregnancy cohorts. Of women with VB-affected vs VB-unaffected pregnancies, 29.0% vs 35.0% had higher education, 13.0% vs 4.9% had a previous record of VB diagnosis irrespective of pregnancy outcome, 6.2% vs 2.1% had a history of a previous VB episode within 20 gestational weeks of a pregnancy ending in a delivery, 22.0% vs 12.0% had at least one previous miscarriage; 19.0% vs 15.0% were smokers (Table 1).

Pregnancies ending in a termination were more likely to be among women who were younger (<24 years), unmarried, nulliparous and unemployed, had a higher prevalence of psychiatric conditions history, a lower level of the highest attained education and were in the first income quartile compared with pregnancies ending in a delivery or miscarriage (Table 1). Pregnancies ending in a miscarriage were more likely than VB-affected pregnancies to be of women older than 35 years and 15.0% had a previous miscarriage (Table 1).

### Cumulative incidences and incidence rates of the cancer outcomes

The median (25^th^-75^th^ percentile) follow-up was 12.6 (6.9-18.2) years. At the end of the follow-up, there were 1 320 cancer events following VB-affected pregnancy, 40 420 events following VB-unaffected pregnancy, 10 300 events following a termination and 4 790 events following a miscarriage (Table 2). At the end of the 24-year follow-up, the cumulative incidence of any cancer was 7.4% following VB-affected pregnancy and 7.5% following VB-unaffected pregnancy, 7.3% following termination and 7.9% following a miscarriage (Figure 1 in the electronic supplementary material). The cumulative incidence of groups of site-specific cancers was also similar between the investigated cohorts. Cancers classified as hormone-related were the most prevalent group with cumulative incidences at the end of follow-up of 3.2%, 3.0%, 2.6%, and 3.0% following VB-affected pregnancy, VB-unaffected pregnancy, termination, and miscarriage, respectively.

**Table 2.**
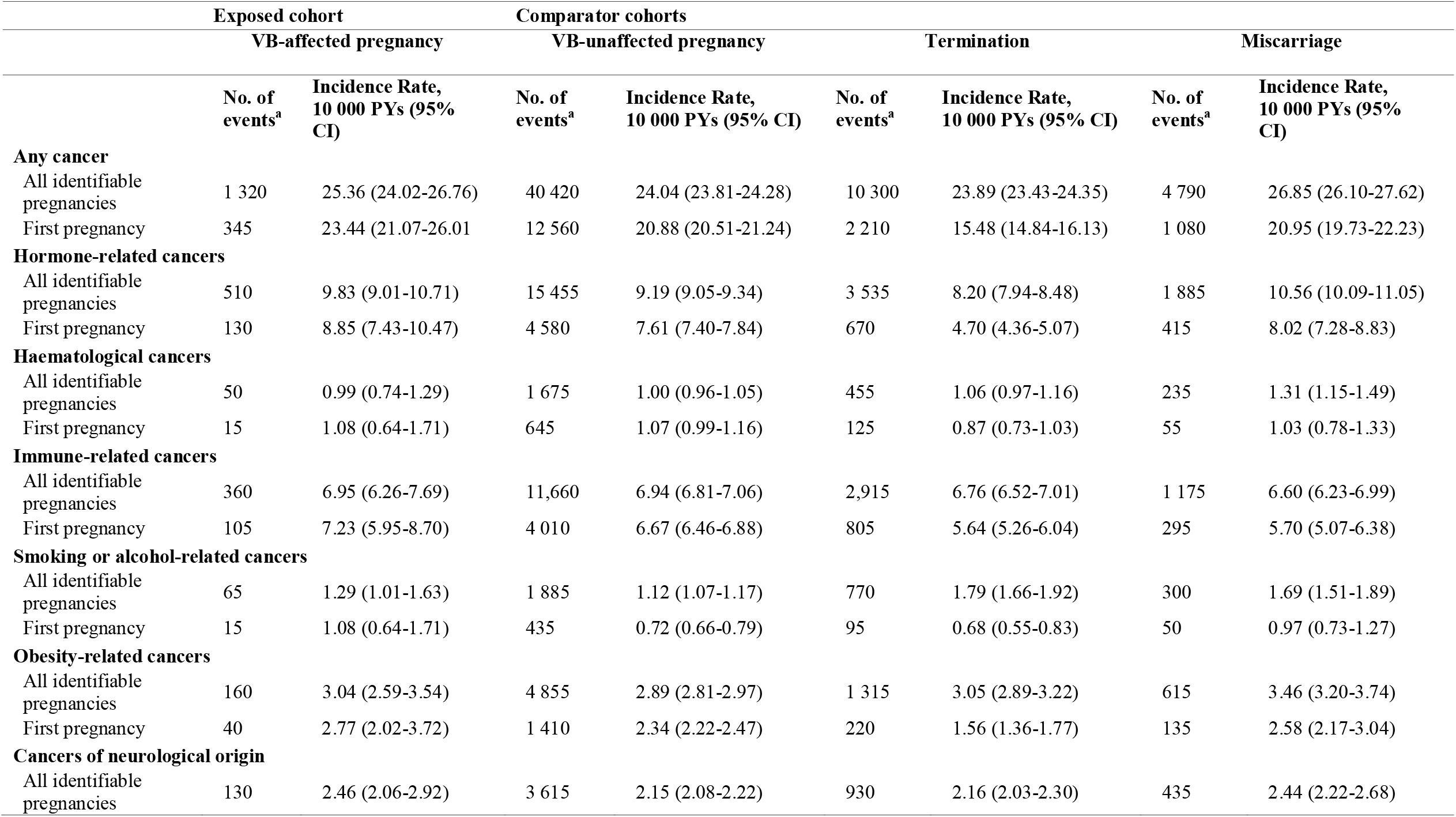
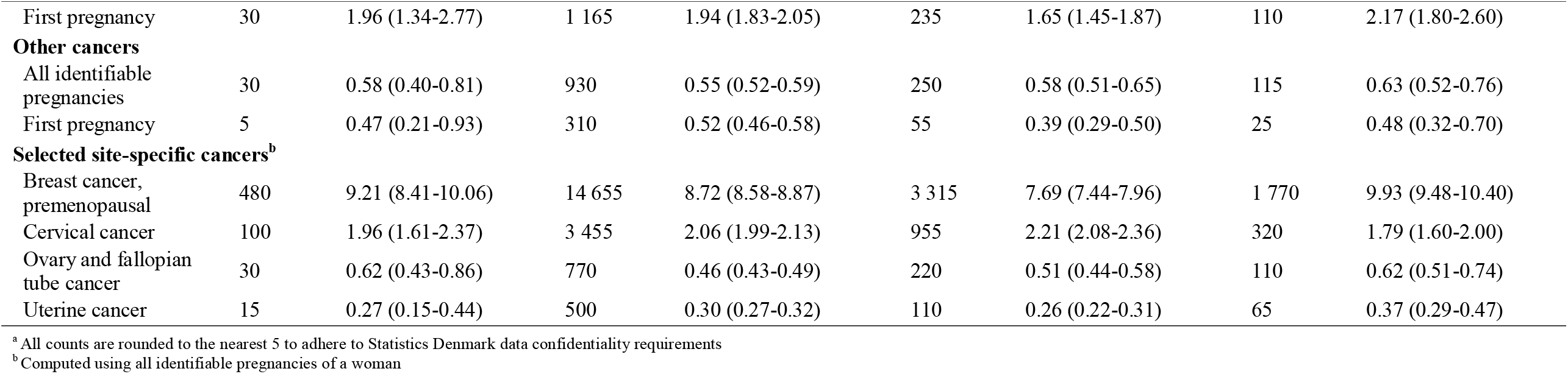
Incidence rates of cancer following vaginal bleeding-affected pregnancy and comparator cohorts, 1995-2018.

|  | Exposed cohort |  | Comparator cohorts |  |  |  |  |  |
| --- | --- | --- | --- | --- | --- | --- | --- | --- |
|  | VB-affected pregnancy |  | VB-unaffected pregnancy |  | Termination |  | Miscarriage |  |
|  | No. of events<sup>a</sup> | Incidence Rate, 10 000 PYs (95% CI) | No. of events<sup>a</sup> | Incidence Rate, 10 000 PYs (95% CI) | No. of events<sup>a</sup> | Incidence Rate, 10 000 PYs (95% CI) | No. of events<sup>a</sup> | Incidence Rate, 10 000 PYs (95% CI) |
| **Any cancer** |  |  |  |  |  |  |  |  |
| All identifiable pregnancies | 1 320 | 25.36 (24.02-26.76) | 40 420 | 24.04 (23.81-24.28) | 10 300 | 23.89 (23.43-24.35) | 4 790 | 26.85 (26.10-27.62) |
| First pregnancy | 345 | 23.44 (21.07-26.01) | 12 560 | 20.88 (20.51-21.24) | 2 210 | 15.48 (14.84-16.13) | 1 080 | 20.95 (19.73-22.23) |
| **Hormone-related cancers** |  |  |  |  |  |  |  |  |
| All identifiable pregnancies | 510 | 9.83 (9.01-10.71) | 15 455 | 9.19 (9.05-9.34) | 3 535 | 8.20 (7.94-8.48) | 1 885 | 10.56 (10.09-11.05) |
| First pregnancy | 130 | 8.85 (7.43-10.47) | 4 580 | 7.61 (7.40-7.84) | 670 | 4.70 (4.36-5.07) | 415 | 8.02 (7.28-8.83) |
| **Haematological cancers** |  |  |  |  |  |  |  |  |
| All identifiable pregnancies | 50 | 0.99 (0.74-1.29) | 1 675 | 1.00 (0.96-1.05) | 455 | 1.06 (0.97-1.16) | 235 | 1.31 (1.15-1.49) |
| First pregnancy | 15 | 1.08 (0.64-1.71) | 645 | 1.07 (0.99-1.16) | 125 | 0.87 (0.73-1.03) | 55 | 1.03 (0.78-1.33) |
| **Immune-related cancers** |  |  |  |  |  |  |  |  |
| All identifiable pregnancies | 360 | 6.95 (6.26-7.69) | 11,660 | 6.94 (6.81-7.06) | 2,915 | 6.76 (6.52-7.01) | 1 175 | 6.60 (6.23-6.99) |
| First pregnancy | 105 | 7.23 (5.95-8.70) | 4 010 | 6.67 (6.46-6.88) | 805 | 5.64 (5.26-6.04) | 295 | 5.70 (5.07-6.38) |
| **Smoking or alcohol-related cancers** |  |  |  |  |  |  |  |  |
| All identifiable pregnancies | 65 | 1.29 (1.01-1.63) | 1 885 | 1.12 (1.07-1.17) | 770 | 1.79 (1.66-1.92) | 300 | 1.69 (1.51-1.89) |
| First pregnancy | 15 | 1.08 (0.64-1.71) | 435 | 0.72 (0.66-0.79) | 95 | 0.68 (0.55-0.83) | 50 | 0.97 (0.73-1.27) |
| **Obesity-related cancers** |  |  |  |  |  |  |  |  |
| All identifiable pregnancies | 160 | 3.04 (2.59-3.54) | 4 855 | 2.89 (2.81-2.97) | 1 315 | 3.05 (2.89-3.22) | 615 | 3.46 (3.20-3.74) |
| First pregnancy | 40 | 2.77 (2.02-3.72) | 1 410 | 2.34 (2.22-2.47) | 220 | 1.56 (1.36-1.77) | 135 | 2.58 (2.17-3.04) |
| **Cancers of neurological origin** |  |  |  |  |  |  |  |  |
| All identifiable pregnancies | 130 | 2.46 (2.06-2.92) | 3 615 | 2.15 (2.08-2.22) | 930 | 2.16 (2.03-2.30) | 435 | 2.44 (2.22-2.68) |
|  | VB-affected pregnancy |  | VB-unaffected pregnancy |  | Termination |  | Miscarriage |  |
| First pregnancy | 30 | 1.96 (1.34-2.77) | 1 165 | 1.94 (1.83-2.05) | 235 | 1.65 (1.45-1.87) | 110 | 2.17 (1.80-2.60) |
| **Other cancers** |  |  |  |  |  |  |  |  |
| All identifiable pregnancies | 30 | 0.58 (0.40-0.81) | 930 | 0.55 (0.52-0.59) | 250 | 0.58 (0.51-0.65) | 115 | 0.63 (0.52-0.76) |
| First pregnancy | 5 | 0.47 (0.21-0.93) | 310 | 0.52 (0.46-0.58) | 55 | 0.39 (0.29-0.50) | 25 | 0.48 (0.32-0.70) |
| **Selected site-specific cancers<sup>b</sup>** |  |  |  |  |  |  |  |  |
| Breast cancer, premenopausal | 480 | 9.21 (8.41-10.06) | 14 655 | 8.72 (8.58-8.87) | 3 315 | 7.69 (7.44-7.96) | 1 770 | 9.93 (9.48-10.40) |
| Cervical cancer | 100 | 1.96 (1.61-2.37) | 3 455 | 2.06 (1.99-2.13) | 955 | 2.21 (2.08-2.36) | 320 | 1.79 (1.60-2.00) |
| Ovary and fallopian tube cancer | 30 | 0.62 (0.43-0.86) | 770 | 0.46 (0.43-0.49) | 220 | 0.51 (0.44-0.58) | 110 | 0.62 (0.51-0.74) |
| Uterine cancer | 15 | 0.27 (0.15-0.44) | 500 | 0.30 (0.27-0.32) | 110 | 0.26 (0.22-0.31) | 65 | 0.37 (0.29-0.47) |

<sup>a</sup> All counts are rounded to the nearest 5 to adhere to Statistics Denmark data confidentiality requirements
<sup>b</sup> Computed using all identifiable pregnancies of a woman

The incidence rate per 10 000 PYs of any cancer was 25.36 (95% CI: 24.02-26.76) for VB-affected pregnancies and 24.04 (95% CI: (23.81-24.28) for VB-affected pregnancies (Table 2). The incidence rates per 10 000 PYs of hormone-related cancers were 9.83 (9.01-10.71) for VB-unaffected pregnancies, 9.19 (9.05-9.34) for VB-affected pregnancies, 8.20 (7.94-8.48) for terminations, and 10.56 (10.09-11.05) for miscarriages. Similarly, the incidence rates per 10 000 PYs of smoking or alcohol-related cancers and obesity-related cancers were slightly higher following VB-affected than VB-unaffected pregnancy. The incidence rates of premenopausal breast cancer, cervical cancer, ovary and fallopian tube cancer, uterine cancer, and cancers at other sites were similar for all investigated cohorts (Table 2 and Table 2 in the electronic supplementary material).

### Hazard ratios for the cancer outcomes

The HR for any cancer in the VB-affected pregnancy cohort was 1.03 (95% CI: 0.97-1.08) vs VB-unaffected pregnancy cohort, 1.03 (95% CI: 0.97-1.09) vs termination cohort, and 0.90 (95% CI: 0.84-0.95) vs miscarriage cohort. In the analyses including the first pregnancy of a woman, the adjusted models yielded nearly identical results as analyses including all identifiable pregnancies of a woman (Table 3).

**Table 3.**
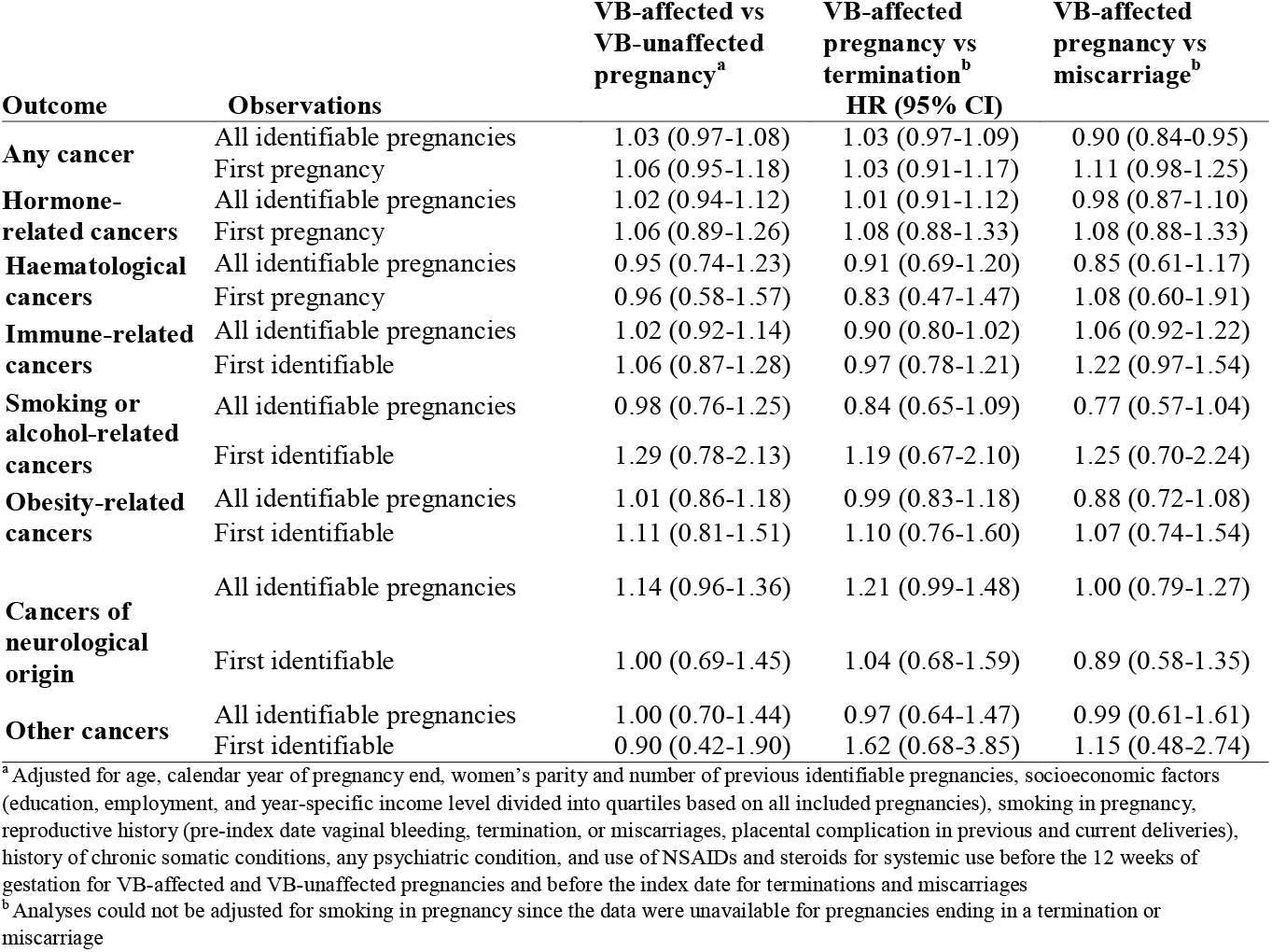
**Hazard ratios for any cancer and cancer groups contrasting VB-affected pregnancy with comparator cohorts (VB-unaffected pregnancy, pregnancies ending in termination or miscarriage), Denmark, 1995-2018**

| Outcome | Observations | VB-affected vs<br>VB-unaffected<br>pregnancy<sup>a</sup> | VB-affected<br>pregnancy vs<br>termination<sup>b</sup><br>HR (95% CI) | VB-affected<br>pregnancy vs<br>miscarriage<sup>b</sup> |
| --- | --- | --- | --- | --- |
| **Any cancer** | All identifiable pregnancies | 1.03 (0.97-1.08) | 1.03 (0.97-1.09) | 0.90 (0.84-0.95) |
|  | First pregnancy | 1.06 (0.95-1.18) | 1.03 (0.91-1.17) | 1.11 (0.98-1.25) |
| **Hormone-related cancers** | All identifiable pregnancies | 1.02 (0.94-1.12) | 1.01 (0.91-1.12) | 0.98 (0.87-1.10) |
|  | First pregnancy | 1.06 (0.89-1.26) | 1.08 (0.88-1.33) | 1.08 (0.88-1.33) |
| **Haematological cancers** | All identifiable pregnancies | 0.95 (0.74-1.23) | 0.91 (0.69-1.20) | 0.85 (0.61-1.17) |
|  | First pregnancy | 0.96 (0.58-1.57) | 0.83 (0.47-1.47) | 1.08 (0.60-1.91) |
| **Immune-related cancers** | All identifiable pregnancies | 1.02 (0.92-1.14) | 0.90 (0.80-1.02) | 1.06 (0.92-1.22) |
|  | First identifiable | 1.06 (0.87-1.28) | 0.97 (0.78-1.21) | 1.22 (0.97-1.54) |
| **Smoking or alcohol-related cancers** | All identifiable pregnancies | 0.98 (0.76-1.25) | 0.84 (0.65-1.09) | 0.77 (0.57-1.04) |
|  | First identifiable | 1.29 (0.78-2.13) | 1.19 (0.67-2.10) | 1.25 (0.70-2.24) |
| **Obesity-related cancers** | All identifiable pregnancies | 1.01 (0.86-1.18) | 0.99 (0.83-1.18) | 0.88 (0.72-1.08) |
|  | First identifiable | 1.11 (0.81-1.51) | 1.10 (0.76-1.60) | 1.07 (0.74-1.54) |
| **Cancers of neurological origin** | All identifiable pregnancies | 1.14 (0.96-1.36) | 1.21 (0.99-1.48) | 1.00 (0.79-1.27) |
|  | First identifiable | 1.00 (0.69-1.45) | 1.04 (0.68-1.59) | 0.89 (0.58-1.35) |
| **Other cancers** | All identifiable pregnancies | 1.00 (0.70-1.44) | 0.97 (0.64-1.47) | 0.99 (0.61-1.61) |
|  | First identifiable | 0.90 (0.42-1.90) | 1.62 (0.68-3.85) | 1.15 (0.48-2.74) |

<sup>a</sup> Adjusted for age, calendar year of pregnancy end, women's parity and number of previous identifiable pregnancies, socioeconomic factors (education, employment, and year-specific income level divided into quartiles based on all included pregnancies), smoking in pregnancy, reproductive history (pre-index date vaginal bleeding, termination, or miscarriages, placental complication in previous and current deliveries), history of chronic somatic conditions, any psychiatric condition, and use of NSAIDs and steroids for systemic use before the 12 weeks of gestation for VB-affected and VB-unaffected pregnancies and before the index date for terminations and miscarriages
<sup>b</sup> Analyses could not be adjusted for smoking in pregnancy since the data were unavailable for pregnancies ending in a termination or miscarriage

There was no strong evidence for an association between VB-affected pregnancy and subsequent risk of hormone-related, haematological, immune-related, obesity-related, cancers of neurologic origin, or other cancers across all contrasts (Table 3). When investigating the risk of smoking or alcohol-related cancers following VB-affected vs VB-unaffected pregnancy the HR was 0.98 (95% CI: 0.79-1.22).

However, when following a first identifiable VB-affected vs VB-unaffected pregnancy of a woman, we observed 1.3-fold increased hazard of smoking or alcohol-related cancers (HR: 1.29, 95% CI: 0.78-2.13) (Table 3). For contrasts of VB-affected pregnancy ending in childbirth vs pregnancy ending in termination or miscarriage, the HRs for smoking or alcohol-related cancers were consistent with slightly reduced risk to no association (HR: 0.84, 95% CI: 0.65-1.09 and HR: 0.77, 95% CI: 0.57-1.04, respectively) in analyses of all identifiable pregnancies. The latter associations could not be adjusted for women’s smoking status, since this information was only available for pregnancies ending in a delivery.

As compared with having VB-unaffected pregnancy, there was no evidence of an increased risk of premenopausal breast cancer (HR: 1.01, 95% CI: 0.92-1.11), cervical cancer (0.95, 0.78-1.16), ovary and fallopian tube cancer (1.20, 0.84-1.71), and uterine cancer (0.85, 0.49-1.42) following VB-affected pregnancy (Table 4). Similarly, we found no association with other investigated site-specific cancers across comparisons with VB-unaffected pregnancy cohort and cohorts of pregnancies ending in a termination or miscarriage (Table 3 in the electronic supplementary material).

**Table 4.**
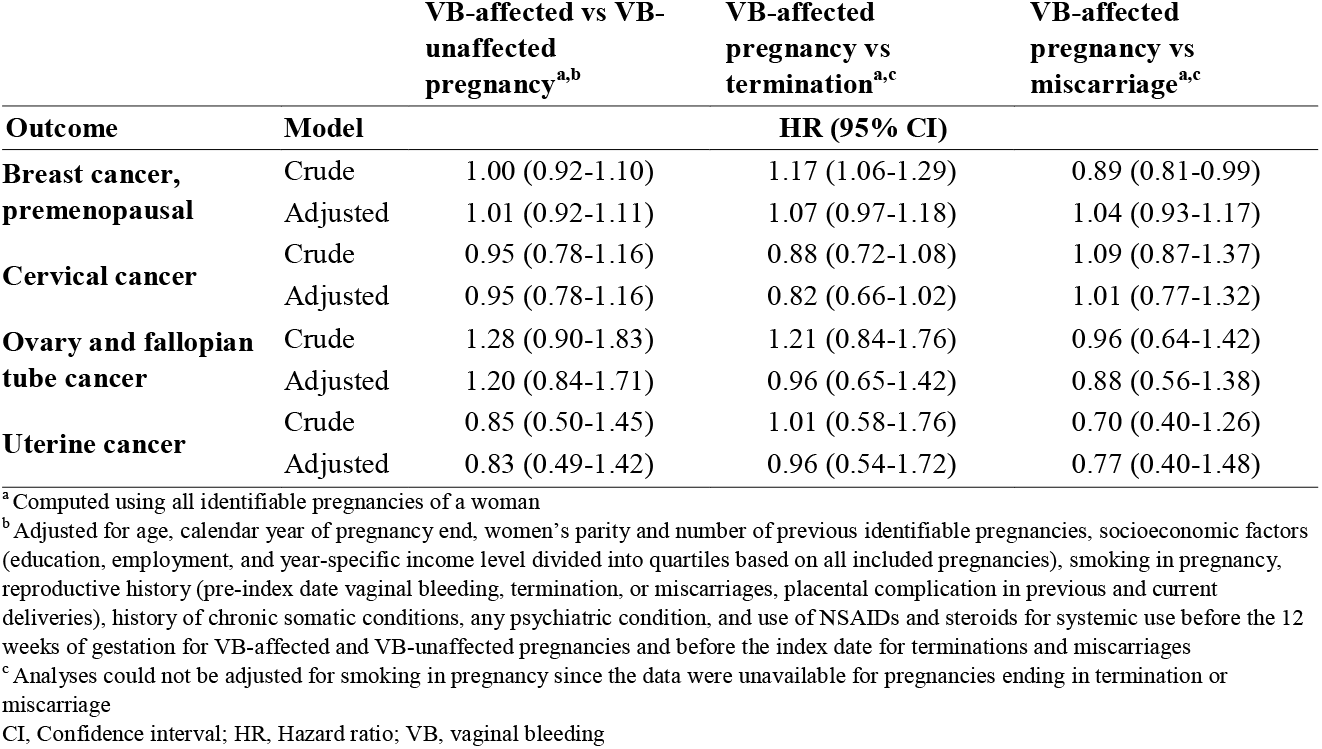
**Hazard ratios for breast cancer, cervical cancer, ovary and fallopian tube cancer, and uterine cancer contrasting VB-affected pregnancy with comparator cohorts, Denmark, 1995-2018**

| Outcome | Model | VB-affected vs VB-unaffected<br>pregnancy<sup>a,b</sup> | VB-affected<br>pregnancy vs<br>termination<sup>a,c</sup> | VB-affected<br>pregnancy vs<br>miscarriage<sup>a,c</sup> |
| --- | --- | --- | --- | --- |
| **Breast cancer,**<br>**premenopausal** | Crude | 1.00 (0.92-1.10) | 1.17 (1.06-1.29) | 0.89 (0.81-0.99) |
|  | Adjusted | 1.01 (0.92-1.11) | 1.07 (0.97-1.18) | 1.04 (0.93-1.17) |
| **Cervical cancer** | Crude | 0.95 (0.78-1.16) | 0.88 (0.72-1.08) | 1.09 (0.87-1.37) |
|  | Adjusted | 0.95 (0.78-1.16) | 0.82 (0.66-1.02) | 1.01 (0.77-1.32) |
| **Ovary and fallopian**<br>**tube cancer** | Crude | 1.28 (0.90-1.83) | 1.21 (0.84-1.76) | 0.96 (0.64-1.42) |
|  | Adjusted | 1.20 (0.84-1.71) | 0.96 (0.65-1.42) | 0.88 (0.56-1.38) |
| **Uterine cancer** | Crude | 0.85 (0.50-1.45) | 1.01 (0.58-1.76) | 0.70 (0.40-1.26) |
|  | Adjusted | 0.83 (0.49-1.42) | 0.96 (0.54-1.72) | 0.77 (0.40-1.48) |

<sup>a</sup> Computed using all identifiable pregnancies of a woman
<sup>b</sup> Adjusted for age, calendar year of pregnancy end, women's parity and number of previous identifiable pregnancies, socioeconomic factors (education, employment, and year-specific income level divided into quartiles based on all included pregnancies), smoking in pregnancy, reproductive history (pre-index date vaginal bleeding, termination, or miscarriages, placental complication in previous and current deliveries), history of chronic somatic conditions, any psychiatric condition, and use of NSAIDs and steroids for systemic use before the 12 weeks of gestation for VB-affected and VB-unaffected pregnancies and before the index date for terminations and miscarriages
<sup>c</sup> Analyses could not be adjusted for smoking in pregnancy since the data were unavailable for pregnancies ending in termination or miscarriage
CI, Confidence interval; HR, Hazard ratio; VB, vaginal bleeding

Sensitivity analysis of pregnancies ending in delivery from 2004 onwards with additional adjustment for woman’s BMI in pregnancy showed results generally consistent with the findings from the main analyses. The HR for smoking or alcohol-related cancers following a first identifiable VB-affected vs VB-unaffected pregnancy of a woman shifted below the null value (HR: 0.50, 95% CI: 0.19-1.33); however, the estimate should be interpreted with caution since it was based on few cancer events and, thus, was imprecise (Table 4 in the electronic supplementary material). HRs for other cancers at specific sites are available in Table 5 in the electronic supplementary material. There was no increased risk of any cancer in analyses of women having their last identifiable pregnancy at the age of 40 years or later (Table 6 in the electronic supplementary material).

## DISCUSSION

### Principal findings

In this registry-based cohort study, we found no evidence of an association between VB in pregnancy carried to delivery and subsequent risk of cancer. When compared with having VB-unaffected pregnancy, there was no increased risk of any cancer and specifically breast, cervical, ovary and fallopian tube, and uterine cancer in women having VB-affected pregnancy. Analysis of a first identifiable VB-affected vs VB-unaffected pregnancy of a woman showed a 1.3-fold increased hazard of smoking or alcohol-related cancers; however, this association attenuated towards the null value in a sensitivity analysis. Similarly, we found no evidence of an association with any cancer or site-specific cancers in women when contrasting having VB-affected pregnancy with termination or miscarriage.

### Comparison with other studies

This study is in line with a previous Danish study reporting no association between VB in pregnancy irrespective of pregnancy end and occult cancer (54). The results of this study also indirectly support the findings of previous research showing that the risk of breast, ovarian and endometrial cancer was not increased in women with a gestation duration of at least 33 weeks (1,2,27).

### Strengths and limitations

Due to nearly complete administrative follow-up for every woman, selection bias at the inclusion of pregnancies in the study is negligible, but potential selection bias from unidentified early miscarriages cannot be ruled out. In the analyses, we controlled for multiple potential confounding factors. Data on proxies of the lifestyle such as smoking and BMI were available only for pregnancies ending in childbirth; moreover, data on BMI was only available for a data subset starting from 2004 and data on in-vitro fertilization or subfertility status were unavailable. The 1.3-fold increased hazard of smoking or alcohol-related cancers in the contrast of a first identifiable VB-affected vs VB-unaffected pregnancy of a woman can potentially be explained by residual confounding by lifestyle factors such as diet, physical activity, and BMI. At the same time, no lifestyle factor of women with VB in pregnancy is likely to dilute the associations and, thus, is unlikely to explain the null associations in analyses contrasting VB-affected pregnancy ending in delivery with having a termination or miscarriage.

Records of VB diagnosis in pregnancy due to threatened miscarriage were not validated in the DNPR. The validity of recording miscarriage diagnoses and procedures for pregnancy termination in the DNPR is high (positive predictive value >90%) (37,55). We could potentially miss recurrent miscarriages within 180 days of each other. However, owing to high specificity of miscarriage diagnoses and few false positives, the misclassification of miscarriages is not a major concern. A diagnostic work-up for threatened miscarriage includes an ultrasound for establishing pregnancy viability, foetal heartbeat, and to eliminate other causes of VB such as ectopic pregnancy (8,56). The prevalence of VB in this study is low (<3%) compared with estimates reported elsewhere (up to 30%) (8–12). We argue that identified VB-affected pregnancies in this study had high specificity, however, a sizeable proportion of VB episodes not resulting in a hospital-based encounter were missed. This may cause bias only if captured episodes of VB systematically differ from all such episodes.

Owing to compulsory reporting of incident primary tumours, the accuracy of cancer diagnoses in the DCR and its coverage is very high, with nearly 90% of tumours having a morphological verification (41). Thus, misclassification of the outcome is unlikely in this study and is not expected to explain the null findings. We did not censor women who would no longer be at risk of endometrial, cervical, and uterine cancers due to hysterectomy (57,58) and the risks of these cancer outcomes may be overestimated.

## CONCLUSION

We found no strong evidence of an association between VB in pregnancy and subsequent risk of cancer during up to 24 years of follow-up.

## Supporting information

Electronic supplementary material

## Data Availability

Data is not available for sharing to protect the identity of participants according to Danish legislation.

## Statements & Declarations

### Author contributions

ED, EHP, HTS, and VE designed the study. HTS facilitated the data application. ED prepared the statistical analysis plan and conducted statistical analyses. EHP provided expertise and supervision of the statistical analyses. All authors participated in the discussion of the results and their interpretation. ED drafted the manuscript, organised the writing, and produced a graphical presentation of the results. HTS provided clinical expertise. All authors equally participated in the critical revision of the manuscript and approved its final version. HTS is the guarantor.

### Ethics approval and consent to participate

No patient consent or ethical approval is required for registry-based research according to Danish legislation. This study was reported to the Danish Data Protection Agency through registration at Aarhus University (record number: AU-2016-051-000001, sequential number 605).

### Consent for publication

Not applicable.

### Data availability

The pseudonymised data used in this study are stored on and accessed via the secure governmental servers of Denmark Statistics (https://www.dst.dk/). According to Danish legislation, Danish research environments may be granted access to data at an individual level. For legal reasons and to protect individuals, whose data were analysed, the dataset used in this study will not be made public.

