## Supplementary material for "Association between having a pregnancy complicated by vaginal bleeding and risk of cancer": Electronic supplementary material

**Supplemental materials**

### **Table 1 Definitions of the exposure, outcomes, and covariables (ICD-8, ICD-10, and ATC codes)**

|  | ICD-8 codes | ICD-10 codes | Procedure codes | ATC codes |
| --- | --- | --- | --- | --- |
| **Exposure and comparators** | | | | |
| Vaginal bleeding in pregnancy (threatened abortion) | 632.39 | DO20.0 | N/A | N/A |
| Miscarriage (spontaneous abortion) | 643xx; 634xx;  645xx | O03; O021.A | N/A | N/A |
| Pregnancy termination (medical or surgical abortion) | 640xx; 641xx; 642xx | O04 | 94520 (abortus provocatus medicamentalis); KLCH (termination of pregnancy) | N/A |
| **Exclusion criteria** |  |  |  |  |
| Any cancer before the index date | 140-209 | C00-C96 | N/A | N/A |
| AIDS before the index date | 07983 | B21-24 | N/A | N/A |
| **Covariables** | | | | |
| Obesity | 277.99 | E66 | N/A | N/A |
| Polycistic ovary syndrome (PCOS) | 256.90 | E28.2 | N/A | N/A |
| Thyroid disorders | 240xx-246xx | E00-E07 | N/A | N/A |
| Conective tissue diseases (CTD) incl. conditions below: | | | N/A | N/A |
| Rheumatic disorders with the heart involvement | 390.99, 391.99, 392.99, 392.09, 393.00, 393.01 | I00.0-I02.9; I05.0-I09.9 | N/A | N/A |
| Rheumatoid arthritis | 712.09-39, 712.59 | M05, M06, M08.0, M08.2-08.9 | N/A | N/A |
| Dermatopolymyositis | 716.09, 716.19 | M33 | N/A | N/A |
| Systemic sclerosis | 734.00-734.09 | M34 | N/A | N/A |
| Systemic lupus erythematosus | 734.19 | M32 | N/A | N/A |
| Sjögren syndrome | 734.90 | M35.0 | N/A | N/A |
| Polyarteritis nodosa | 446.09 | M30.0 | N/A | N/A |
| Wegener granulomatosis | 446.29 | M31.3 | N/A | N/A |
| Temporal arteritis and polymyalgia rheumatica | 46.30-446.39 | M31.5, M31.6, M35.3 | N/A | N/A |
| Psoriatic arthritis | 696.09 | L40.5, M07.0-07.3 | N/A | N/A |
| Ankylosing spondylitis | 712.49 | M45.9, M08.1 | N/A | N/A |
| Arthritis not further specified | 715.99 | M13.0, M13.1, M13.8, M13.9 | N/A | N/A |
| Unspecified rheumatism (including fibromyalgia and myalgia) | 717.90, 717.99, 718.99 | M25.5, M25.6, M25.8, M25.9, M62.6, M62.8, M62.9, M79.0, M79.1, M79.8, M79.9 | N/A | N/A |
| Localized scleroderma | 701.01-701.09 | L94.0-L94.3 | N/A | N/A |
| Localized (discoid) lupus erythematosus | 695.49 | L93.0-L93.2 | N/A | N/A |
| CTD not further specified | 734.91, 734.99 | M35.1, M35.2, M35.4-M35.9, M79.3 | N/A | N/A |
| Sarcoidosis | 135.99 | D86 | N/A | N/A |
| Amyloidosis | 276.00-276.09 | E85 | N/A | N/A |
| COPD | 490-493; 515-518 | J40-J47; J60-J67;  J68.4; J70.1; J70.3;  J84.1; J92.0; J96.1;  J98.2; J98.3 | N/A | N/A |
| Chronic kidney diseases | 249.02, 250.02, 753.10-753.19, 582, 583, 584, 590.09, 593.20, 792 | 249.02, 250.02, 753.10-753.19, 582, 583, 584, 590.09, 593.20, 792;  E10.2, E11.2, E14.2,  N03, N05, N11.0,  N14; N16, N18-N19,  N26.9, Q61.1-Q61.4 | N/A | N/A |
| Chronic liver diseases | 571 | K70.0, K70.3, K71.7,  K73, K74, K76.0,  B18, I85 | N/A | N/A |
| Preeclampsia superimposed on chronic hypertension | N/A | O11 | N/A | N/A |
| Gestational proteinuria | 637.01, 637.02 | O12 | N/A | N/A |
| Gestational hypertension | 637.00 | O13 | N/A | N/A |
| Preeclampsia | 637.03, 637.04, 637.09 | O14 | N/A | N/A |
| Eclampsia | 637.19 | O15 | N/A | N/A |
| Any hypertensive disorder of pregnancy | 637.01, 637.02, 637.00, 637.03, 637.04, 637.09, 637.19 | O11- O15 | N/A | N/A |
| Placenta praevia | 651.0x; 651.1x; 651.2x; 651.3x | O44 | N/A | N/A |
| Abruptio placentae | 6321x; 6514x | O45 | N/A | N/A |
| Gestational diabetes | 634.74, 644.9x, Y6449 | O24.4, O24.9 | N/A | N/A |
| Hypercholesterolemia or hyperlipidemia | 272.01, 272.08, 272.09,  272.00 | E78.1-E78.5,  E78.0 | N/A | C10AA, C10B |
| Diabetes mellitus (same codes are used for diabetes mellhistoryitus type 1 and 2 until 1987) | 249.0-250.9 | E10-E14, E89.1, G59.0, G63.2, G73.0A, G99.0C, H28.0, H360, I792.A, M14.2, N08.3 | N/A | A10A, A10B |
| Hypertension | 400.09-404.99 | I10.0-I15.9 | N/A | C02A, C02B, C02C (alpha-blockers); C02DA, C02L, C03A, C03B, C03D, C03E, C03X, C07C, C07D, C08G, C09BA, C09DA, C09XA52 (non-loop diuretics); C02BD, C02DD, C02DG, C04, C05 (vasodilators); C07 (beta-blockers); C08, C07F, C09BB, C09DB (calcium channel blockers); C09 (RAS-acting agents: ACE inhibitors and ARBs). Threatment was classified as antihypertensive therapy when 2+ distinct antihypertensives classes were dispensed within 180 days of each other. |
| **Any psychiatric disorder** | 290-315 | F00-F99 | N/A | Psychotropic medications (N05A [antipsychotics], N06 [[psychoanaleptics](https://www.whocc.no/atc_ddd_index/?code=N06&showdescription=no)]) |
| Alcohol or drug abuse | 291x, 303x9, 303.20, 303.28, 303.90, 294.39, 304x9; 577.10, 571.09, 571.10 | F10, F11-F19; Z72.1, T51.0, K86.0, G31.2, G62.1, G72.1, I42.6, K29.2, R78.0, Z71.4 | N/A | N/A |
| **Outcomes** | | | | |
| ***Obesity-related cancers*** |  |  |  |  |
| Esophagus* | - | C15 | N/A | N/A |
| Pancreas | - | C25 | N/A | N/A |
| Colon including the rectosigmoid junction | - | C18-C19 | N/A | N/A |
| Rectum | - | C20 | N/A | N/A |
| Breast, postmenopausal (≥60 years) | - | C50 |  |  |
| Uterus | - | C54-C55 | N/A | N/A |
| Kidney | - | C64 | N/A | N/A |
| Gallbladder and bile ducts | - | C23-C24 | N/A | N/A |
| Thyroid gland | - | C73 | N/A | N/A |
| ***Smoking- or alcohol-related cancers*** |  |  |  |  |
| Lip* | - | C00 | N/A | N/A |
| Tongue | - | C01-02 | N/A | N/A |
| Mouth | - | C03-06 | N/A | N/A |
| Tonsils and pharynx | - | C09-C13 | N/A | N/A |
| Other and poorly specified locations in lip, oral cavity, and pharynx* | - | C14 | N/A | N/A |
| Larynx* | - | C32 | N/A | N/A |
| Other and poorly specified locations in airways and respiratory organs | - | C39 | N/A | N/A |
| Stomach | - | C16 | N/A | N/A |
| Small intestine | - | C17 | N/A | N/A |
| Liver, including intrahepatic bile ducts | - | C22 | N/A | N/A |
| Lung, bronchus and trachea | - | C33-C34 | N/A | N/A |
| Renal pelvis* | - | C65 | N/A | N/A |
| Ureter* | - | C66 | N/A | N/A |
| Urinary bladder | - | C67 | N/A | N/A |
| ***Hematological cancers*** |  |  |  |  |
| Hodgkin’s lymphoma (included morphologic code 965-966) | - | C81 | N/A | N/A |
| Non-Hodgkin’s lymphoma, excluding leukemia and myelomatosis (including morphologic code 959, 967-972) | - | C82-85 | N/A | N/A |
| Malignant myeloproliferative disease* | - | C88 | N/A | N/A |
| Multiple myeloma and other plasma cell neoplasms | - | C90 | N/A | N/A |
| Myeloid leukemia | - | C92 | N/A | N/A |
| Lymphocytic leukemia | - | C91 | N/A | N/A |
| Monocytic leukemia* | - | C93 | N/A | N/A |
| Other leukemia* | - | C94-95 | N/A | N/A |
| Other and unspecified cancers of lymphoid, hematopoietic, and related tissues* | - | C96 | N/A | N/A |
| Metastasis and unspecified cancer in lymph nodes (only when no primary tumor is coded) | - | C77-79 (only if no primary tumor is coded) | N/A | N/A |
| ***Immune-related cancers*** | - |  | N/A | N/A |
| Anus and anal canal, excluding malignant melanomas (morphologic code 872-879) and basal cell cancers (morphologic code 809) | - | C21 | N/A | N/A |
| Cervix | - | C53 | N/A | N/A |
| External female genitalia, excluding basal cell carcinomas (morphological code 809)* | - | C51 | N/A | N/A |
| Malignant melanoma, including those located in anus and anal canal (morphological code 872-879) | - | C43 | N/A | N/A |
| Non-melanoma skin cancers, excluding basal cell carcinoma (morphologic code 809) | - | C44 | N/A | N/A |
| ***Cancers of neurological origin*** |  |  |  |  |
| Meningioma | - | C70, D32, D42 | N/A | N/A |
| Brain, including hypophysis, corpus pineale, and ductus craniopharyngealis | - | C71, C751-753, D330-D332, D352-D354, D430-D432, D443-D445 | N/A | N/A |
| Spinal cord, cranial nerves, and other parts of central nervous system |  | C72, D333-D339, D433-D439 | N/A | N/A |
| ***Hormone-related cancers*** |  |  |  |  |
| Vagina, excluding basal cell carcinomas (morphological code 809)* | - | C52 | N/A | N/A |
| Breast, premenopausal (<60 years) | - | C50 | N/A | N/A |
| Ovary and fallopian tube | - | C56, C570-574 | N/A | N/A |
| ***All other cancers*** |  |  |  |  |
| Salivary glands | - | C07-08 | N/A | N/A |
| Other and ill-defined cancers of digestive organs* | - | C26 |  |  |
| Nasal cavity, middle ear, and accessory sinuses | - | C30-C31 |  |  |
| Thymus* | - | C37 |  |  |
| Heart and mediastinum* | - | C381-383, C388 | N/A | N/A |
| Pleura, including mesothelioma pleura* | - | C384, C450 | N/A | N/A |
| Bone and articular cartilage | - | C40-C41 | N/A | N/A |
| Kaposi’s sarcoma* | - | C46, B210 | N/A | N/A |
| Mesothelioma* | - | C451-C459 | N/A | N/A |
| Peripheral nerves and autonomic nervous system* | - | C47 | N/A | N/A |
| Retroperitoneum and peritoneum* | - | C48 | N/A | N/A |
| Malignant neoplasm of other connective and soft tissue | - | C49 | N/A | N/A |
| Placenta* | - | C58 | N/A | N/A |
| Other and unspecified cancers in female genital organs | - | C577-579 | N/A | N/A |
| Other and unspecified cancers in urinary organs* | - | C68 | N/A | N/A |
| Eye and adnexa | - | С69 | N/A | N/A |
| Adrenal gland* | - | C74 | N/A | N/A |
| Other endocrine structures, excluding hypophysis, corpus pineale, and ductus craniopharyngealis* | - | C750, C754-759 | N/A | N/A |
| Malignant neoplasm at other, ill-defined, or unspecified sites | - | C76, C80 | N/A | N/A |
| Malignant neoplasms at independent (primary) multiple sites | - | C97 | N/A | N/A |
| Notes: Other types of malignancy, incl. cancer *in-situ,* were not included as end points in this study  * Not included in analyses of individual site-specific cancers due to few accrued (<100) events between 1995 and 2018  Medication prescriptions as part of covariables’ definitions were ascertained as at least one prescription of a drug any time before 12 gestational weeks for VB-affected and VB-unaffected pregnancies and before the index date for terminations and miscarriages | | | | |

### **Figure 1 Cumulative incidence of any cancer at the end of 24-year follow-up period following VB-affected pregnancy cohort and comparator cohorts, Denmark, 1995-2018**

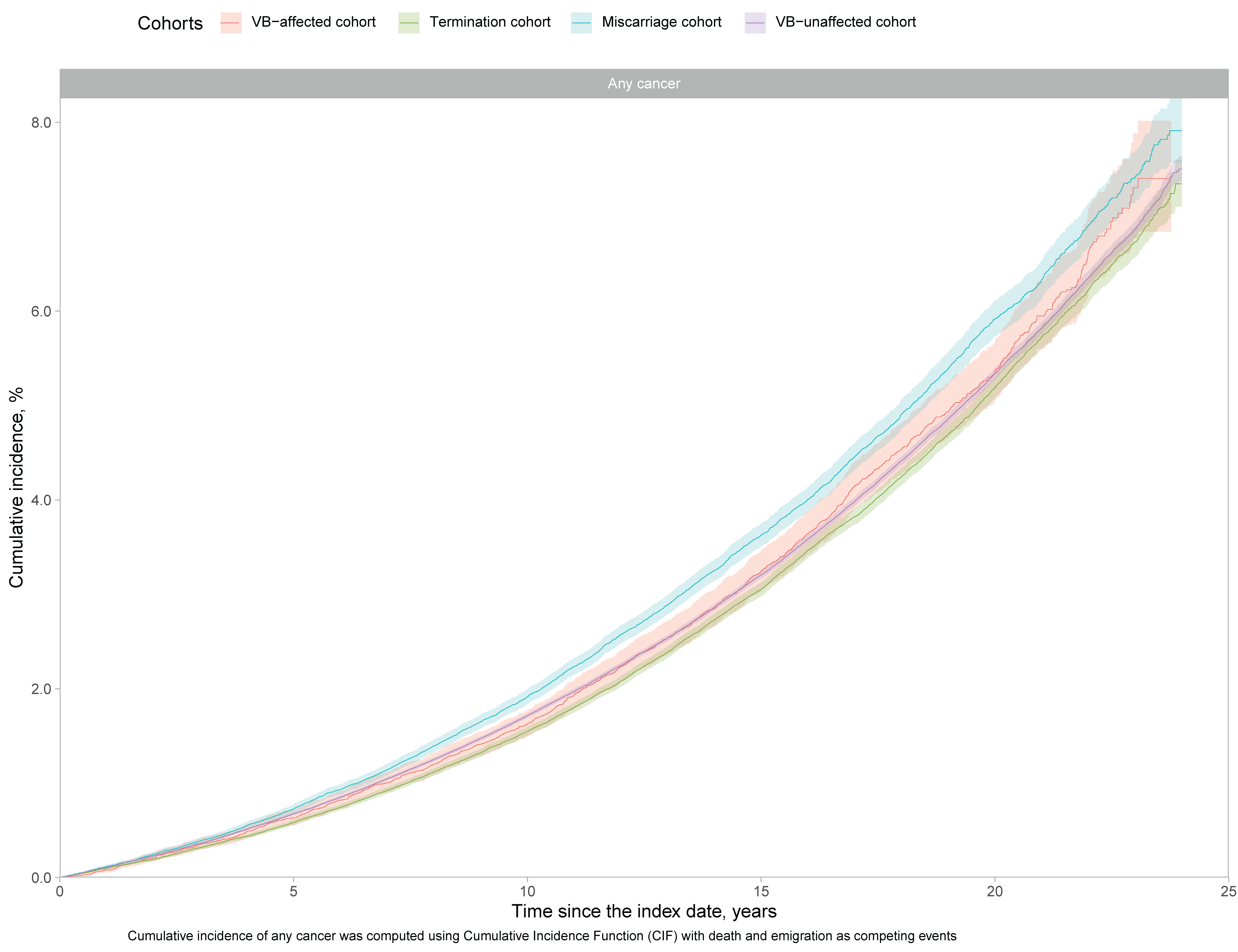

### **Table 2 Number of events and incidence rates of cancer at specific sites, 1995-2018**

| **Cancer site** | **VB-affected pregnancy,**  **N = 37 082** | | **VB-unaffected pregnancy,**  **N = 1 363 614** | | **Termination,**  **N = 324 328** | | **Miscarriage,**  **N = 137 104** | |
| --- | --- | --- | --- | --- | --- | --- | --- | --- |
|  | **N, events**^a^ | **IR (95%) per 10 000 PY** | **N, events**^a^ | **IR (95%) per 10 000 PY** | **N, events**^a^ | **IR (95%) per 10 000 PY** | **N, events**^a^ | **IR (95%) per 10 000 PY** |
| Breast, postmenopausal | 5 | 0.02 (0.00-0.09) | 70 | 0.04 (0.03-0.05) | 85 | 0.19 (0.15-0.24) | 45 | 0.24 (0.18-0.32) |
| Lung, bronchus, trachea | 30 | 0.60 (0.41-0.84) | 1 005 | 0.60 (0.56-0.63) | 490 | 1.14 (1.04-1.24) | 175 | 0.97 (0.83-1.12) |
| Malignant melanoma | 205 | 3.95 (3.43-4.52) | 6 580 | 3.92 (3.82-4.01) | 1 455 | 3.38 (3.21-3.56) | 675 | 3.79 (3.51-4.08) |
| Colon including the rectosigmoid junction | 40 | 0.79 (0.57-1.06) | 1 145 | 0.68 (0.64-0.72) | 350 | 0.82 (0.74-0.91) | 135 | 0.76 (0.64-0.89) |
| Non-melanoma skin cancer excl. basal cell carcinomas | 40 | 0.79 (0.57-1.06) | 1 270 | 0.75 (0.71-0.80) | 370 | 0.86 (0.77-0.95) | 135 | 0.75 (0.63-0.89) |
| Myeloid leukemia | 15 | 0.27 (0.15-0.44) | 335 | 0.20 (0.18-0.22) | 95 | 0.22 (0.18-0.27) | 50 | 0.29 (0.22-0.38) |
| Brain, hypophysis, corpus pineale, ductus craniopharingealis | 70 | 1.39 (1.09-1.74) | 1 765 | 1.05 (1.00-1.10) | 510 | 1.18 (1.08-1.29) | 210 | 1.18 (1.03-1.35) |
| Rectum | 25 | 0.48 (0.32-0.70) | 685 | 0.41 (0.38-0.44) | 180 | 0.41 (0.36-0.48) | 80 | 0.44 (0.35-0.54) |
| Metastasis and unspecifiedcancer in lymph nodes (no primary tumor recorded) | 10 | 0.23 (0.13-0.39) | 335 | 0.20 (0.18-0.22) | 115 | 0.27 (0.23-0.32) | 25 | 0.15 (0.10-0.21) |
| Non-Hodgkin's lymphoma | 10 | 0.23 (0.13-0.39) | 625 | 0.37 (0.34-0.40) | 190 | 0.45 (0.39-0.51) | 80 | 0.45 (0.36-0.56) |
| Meningioma | 40 | 0.75 (0.54-1.02) | 1 230 | 0.73 (0.69-0.77) | 290 | 0.68 (0.60-0.76) | 155 | 0.88 (0.75-1.02) |
| Thyroid gland | 50 | 0.98 (0.74-1.28) | 1 670 | 0.99 (0.95-1.04) | 345 | 0.80 (0.72-0.89) | 185 | 1.05 (0.91-1.21) |
| Pancreas | 5 | 0.13 (0.06-0.26) | 280 | 0.17 (0.15-0.19) | 95 | 0.22 (0.18-0.27) | 35 | 0.19 (0.13-0.26) |
| Multiple myeloma and other plasma cell neoplasms | 5 | 0.13 (0.06-0.26) | 180 | 0.11 (0.09-0.12) | 55 | 0.13 (0.10-0.17) | 30 | 0.17 (0.12-0.24) |
| Kidney | 0 | NA | 0 | NA | 0 | NA | 0 | NA |
| Liver, incl. intrahepatic bile ducts | 0 | NA | 0 | NA | 0 | NA | 0 | NA |
| Eye and adnexa | 0 | NA | 0 | NA | 0 | NA | 0 | NA |
| Spinal cord, cranial nerves and other parts of CNS | 15 | 0.33 (0.20-0.51) | 615 | 0.37 (0.34-0.40) | 130 | 0.31 (0.26-0.36) | 70 | 0.38 (0.30-0.48) |
| Lymphocytic leukemia | 5 | 0.10 (0.04-0.21) | 200 | 0.12 (0.10-0.14) | 45 | 0.10 (0.08-0.14) | 30 | 0.17 (0.12-0.24) |
| Tonsils and pharinx | 5 | 0.10 (0.04-0.21) | 140 | 0.08 (0.07-0.10) | 60 | 0.14 (0.11-0.18) | 20 | 0.12 (0.08-0.18) |
| Mouth | 5 | 0.10 (0.04-0.21) | 95 | 0.06 (0.05-0.07) | 25 | 0.06 (0.04-0.08) | 20 | 0.10 (0.06-0.16) |
| Tongue | 5 | 0.10 (0.04-0.21) | 90 | 0.05 (0.04-0.07) | 25 | 0.06 (0.04-0.08) | 10 | 0.04 (0.02-0.08) |
| Nasal cavity, middle ear and accessory sinuses | 5 | 0.10 (0.04-0.21) | 80 | 0.05 (0.04-0.06) | 25 | 0.05 (0.03-0.08) | 10 | 0.05 (0.02-0.09) |
| Stomach | 10 | 0.19 (0.10-0.34) | 240 | 0.14 (0.13-0.16) | 60 | 0.14 (0.11-0.18) | 30 | 0.17 (0.12-0.24) |
| External female genitalia excl. basal cell carcinomas | 10 | 0.19 (0.10-0.34) | 185 | 0.11 (0.10-0.13) | 75 | 0.17 (0.13-0.21) | 25 | 0.15 (0.10-0.22) |
| Urinary bladder | 5 | 0.06 (0.02-0.15) | 80 | 0.05 (0.04-0.06) | 35 | 0.08 (0.06-0.11) | 15 | 0.07 (0.04-0.12) |
| Anus and anal canal excl. melanomas and basal cell carcinomas | 5 | 0.06 (0.02-0.15) | 175 | 0.10 (0.09-0.12) | 65 | 0.15 (0.11-0.19) | 20 | 0.12 (0.08-0.18) |
| Malignant neoplasm of other connective and soft tissue | 10 | 0.17 (0.09-0.32) | 280 | 0.17 (0.15-0.19) | 70 | 0.17 (0.13-0.21) | 30 | 0.16 (0.11-0.22) |
| Malignant neoplasm of other, ill defined, or unspecified sites | 5 | 0.08 (0.03-0.18) | 80 | 0.05 (0.04-0.06) | 15 | 0.04 (0.02-0.06) | 10 | 0.06 (0.03-0.11) |
| Small intestine | 5 | 0.08 (0.03-0.18) | 100 | 0.06 (0.05-0.07) | 25 | 0.06 (0.04-0.08) | 15 | 0.07 (0.04-0.12) |
| Salivary gland | 5 | 0.08 (0.03-0.18) | 75 | 0.05 (0.04-0.06) | 30 | 0.06 (0.04-0.09) | 10 | 0.07 (0.04-0.11) |
| Hodgkin's lymphoma | 10 | 0.21 (0.11-0.37) | 280 | 0.17 (0.15-0.19) | 55 | 0.13 (0.10-0.16) | 30 | 0.18 (0.13-0.25) |
| Bone and articular cartilage | 5 | 0.04 (0.01-0.12) | 115 | 0.07 (0.06-0.08) | 30 | 0.07 (0.05-0.10) | 20 | 0.10 (0.06-0.16) |

NA=non-available

^a^All counts are rounded to the nearest 5 to adhere to Denmark Statistics data confidentiality requirements

### **Table 3 Hazard ratios with 95% confidence intervals for cancer at specific sites, Denmark, 1995-2018**

|  |  | **VB-affected vs VB-unaffected pregnancy^a,b^** | **VB-affected pregnancy vs termination^a,c^** | **VB-affected pregnancy vs miscarriage^a,c^** |
| --- | --- | --- | --- | --- |
| **Outcome** | **Model** | **HR (95% CI)** | | |
| Anus and anal canal excl. melanomas and basal cell carcinomas | Crude | 0.52 (0.17-1.64) | 0.38 (0.12-1.22) | 0.45 (0.13-1.50) |
|  | Adjusted | 0.48 (0.15-1.52) | 0.43 (0.13-1.39) | 0.79 (0.24-2.64) |
| Bone and articular cartilage | Crude | 0.55 (0.14-2.21) | 0.51 (0.12-2.14) | 0.38 (0.09-1.65) |
|  | Adjusted | 0.58 (0.14-2.35) | 0.42 (0.10-1.82) | 0.38 (0.08-1.92) |
| Brain, hypophysis, corpus pineale, ductus craniopharingealis | Crude | 1.30 (1.02-1.64) | 1.16 (0.91-1.49) | 1.15 (0.88-1.51) |
|  | Adjusted | 1.36 (1.07-1.73) | 1.21 (0.93-1.58) | 1.06 (0.77-1.46) |
| Breast, postmenopausal | Crude | 0.50 (0.07-3.62) | 0.10 (0.01-0.74) | 0.08 (0.01-0.55) |
|  | Adjusted | 0.36 (0.05-2.63) | 0.21 (0.03-1.51) | 0.30 (0.08-1.10) |
| Colon including the rectosigmoid junction | Crude | 1.10 (0.81-1.51) | 0.95 (0.69-1.31) | 1.02 (0.72-1.45) |
|  | Adjusted | 1.09 (0.80-1.50) | 0.94 (0.67-1.32) | 1.00 (0.67-1.50) |
| External female genitalia excl. basal cell carcinomas | Crude | 1.65 (0.88-3.13) | 1.11 (0.57-2.15) | 1.20 (0.58-2.48) |
|  | Adjusted | 1.52 (0.80-2.88) | 1.26 (0.62-2.52) | 1.32 (0.64-2.72) |
| Eye and adnexa | Crude | NA (NA-NA) | NA (NA-NA) | NA (NA-NA) |
|  | Adjusted | NA (NA-NA) | NA (NA-NA) | NA (NA-NA) |
| Hodgkin's lymphoma | Crude | 1.29 (0.71-2.36) | 1.66 (0.87-3.16) | 1.16 (0.59-2.31) |
|  | Adjusted | 1.39 (0.76-2.56) | 1.41 (0.69-2.89) | 1.31 (0.59-2.91) |
| Kidney | Crude | NA (NA-NA) | NA (NA-NA) | NA (NA-NA) |
|  | Adjusted | NA (NA-NA) | NA (NA-NA) | NA (NA-NA) |
| Liver, incl. intrahepatic bile ducts | Crude | NA (NA-NA) | NA (NA-NA) | NA (NA-NA) |
|  | Adjusted | NA (NA-NA) | NA (NA-NA) | NA (NA-NA) |
| Lung, bronchus, trachea | Crude | 0.94 (0.66-1.35) | 0.52 (0.36-0.75) | 0.57 (0.39-0.84) |
|  | Adjusted | 0.80 (0.56-1.14) | 0.63 (0.43-0.91) | 0.62 (0.41-0.95) |
| Lymphocytic leukemia | Crude | 0.77 (0.32-1.86) | 0.90 (0.36-2.27) | 0.53 (0.20-1.36) |
|  | Adjusted | 0.76 (0.31-1.84) | 0.95 (0.36-2.48) | 0.44 (0.15-1.25) |
| Malignant melanoma | Crude | 0.98 (0.86-1.13) | 1.15 (1.00-1.33) | 1.02 (0.87-1.19) |
|  | Adjusted | 1.07 (0.93-1.23) | 1.05 (0.90-1.22) | 1.08 (0.90-1.29) |
| Malignant neoplasm of other connective and soft tissue | Crude | 1.03 (0.53-2.00) | 1.02 (0.51-2.04) | 1.06 (0.50-2.25) |
|  | Adjusted | 0.96 (0.49-1.88) | 0.99 (0.47-2.08) | 1.23 (0.51-2.98) |
| Malignant neoplasm of other, ill defined, or unspecified sites | Crude | 1.56 (0.57-4.25) | 2.04 (0.68-6.11) | 1.22 (0.39-3.82) |
|  | Adjusted | 1.66 (0.60-4.55) | 1.53 (0.48-4.83) | 0.59 (0.15-2.27) |
| Meningioma | Crude | 0.98 (0.71-1.35) | 1.08 (0.77-1.51) | 0.82 (0.58-1.16) |
|  | Adjusted | 0.97 (0.71-1.34) | 1.21 (0.85-1.71) | 0.96 (0.64-1.44) |
| Metastasis and unspecifiedcancer in lymph nodes (no primary tumor recorded) | Crude | 1.16 (0.65-2.06) | 0.85 (0.47-1.54) | 1.56 (0.79-3.09) |
|  | Adjusted | 1.02 (0.57-1.83) | 0.84 (0.45-1.56) | 1.37 (0.61-3.08) |
| Mouth | Crude | 1.60 (0.65-3.93) | 1.66 (0.63-4.35) | 0.94 (0.35-2.53) |
|  | Adjusted | 1.39 (0.56-3.43) | 2.38 (0.84-6.71) | 1.66 (0.55-5.04) |
| Multiple myeloma and other plasma cell neoplasms | Crude | 1.18 (0.56-2.52) | 1.02 (0.47-2.25) | 0.76 (0.33-1.72) |
|  | Adjusted | 1.01 (0.47-2.16) | 1.20 (0.53-2.72) | 1.20 (0.47-3.06) |
| Myeloid leukemia | Crude | 1.32 (0.78-2.26) | 1.22 (0.70-2.15) | 0.90 (0.50-1.63) |
|  | Adjusted | 1.29 (0.75-2.21) | 1.13 (0.62-2.07) | 0.61 (0.30-1.22) |
| Nasal cavity, middle ear and accessory sinuses | Crude | 1.96 (0.79-4.84) | 1.77 (0.67-4.67) | 1.85 (0.62-5.51) |
|  | Adjusted | 2.10 (0.85-5.23) | 2.15 (0.75-6.13) | 2.56 (0.86-7.67) |
| Non-Hodgkin's lymphoma | Crude | 0.59 (0.33-1.05) | 0.51 (0.28-0.91) | 0.49 (0.27-0.90) |
|  | Adjusted | 0.60 (0.34-1.06) | 0.53 (0.29-0.97) | 0.63 (0.32-1.23) |
| Non-melanoma skin cancer excl. basal cell carcinomas | Crude | 1.00 (0.73-1.37) | 0.90 (0.65-1.25) | 1.02 (0.72-1.44) |
|  | Adjusted | 0.99 (0.73-1.36) | 0.79 (0.57-1.11) | 1.15 (0.76-1.73) |
| Pancreas | Crude | 0.77 (0.36-1.63) | 0.62 (0.29-1.35) | 0.69 (0.30-1.56) |
|  | Adjusted | 0.71 (0.33-1.50) | 0.66 (0.30-1.45) | 0.38 (0.15-0.97) |
| Rectum | Crude | 1.12 (0.75-1.67) | 1.16 (0.76-1.76) | 1.05 (0.67-1.65) |
|  | Adjusted | 1.10 (0.73-1.64) | 1.20 (0.77-1.86) | 1.58 (0.94-2.65) |
| Salivary gland | Crude | 1.71 (0.63-4.68) | 1.19 (0.42-3.38) | 1.10 (0.36-3.43) |
|  | Adjusted | 1.54 (0.56-4.26) | 1.13 (0.37-3.42) | 1.17 (0.38-3.62) |
| Small intestine | Crude | 1.27 (0.47-3.45) | 1.32 (0.46-3.78) | 1.05 (0.34-3.21) |
|  | Adjusted | 1.29 (0.47-3.52) | 1.12 (0.37-3.39) | 1.26 (0.41-3.89) |
| Spinal cord, cranial nerves and other parts of CNS | Crude | 0.87 (0.54-1.41) | 1.04 (0.63-1.73) | 0.83 (0.49-1.42) |
|  | Adjusted | 0.90 (0.55-1.45) | 0.92 (0.54-1.56) | 0.81 (0.44-1.49) |
| Stomach | Crude | 1.32 (0.70-2.48) | 1.33 (0.68-2.60) | 1.09 (0.53-2.24) |
|  | Adjusted | 1.32 (0.70-2.50) | 1.28 (0.63-2.60) | 0.98 (0.43-2.26) |
| Thyroid gland | Crude | 0.97 (0.74-1.29) | 1.21 (0.90-1.63) | 0.92 (0.67-1.25) |
|  | Adjusted | 1.01 (0.76-1.33) | 1.11 (0.81-1.53) | 0.96 (0.67-1.39) |
| Tongue | Crude | 1.69 (0.69-4.15) | 1.70 (0.65-4.45) | 1.97 (0.64-6.01) |
|  | Adjusted | 1.58 (0.64-3.92) | 1.80 (0.64-5.04) | 4.98 (1.62-15.31) |
| Tonsils and pharinx | Crude | 1.08 (0.44-2.63) | 0.70 (0.28-1.75) | 0.74 (0.28-1.95) |
|  | Adjusted | 1.04 (0.42-2.54) | 0.78 (0.30-1.99) | 0.71 (0.24-2.12) |
| Urinary bladder | Crude | 1.12 (0.35-3.53) | 0.70 (0.22-2.29) | 0.73 (0.21-2.55) |
|  | Adjusted | 1.01 (0.32-3.23) | 0.87 (0.26-2.94) | 0.40 (0.09-1.68) |
| ^a^ Computed using all identifiable pregnancies of a woman  ^b^ Adjusted for age, calendar year of pregnancy end, women’s parity and number of previous identifiable pregnancies, socioeconomic factors (education, employment, and year-specific income level divided into quartiles based on all included pregnancies), smoking in pregnancy, reproductive history (pre-index date vaginal bleeding, termination, or miscarriages, placental complication in previous deliveries), history of chronic somatic conditions, any psychiatric condition, and use of NSAIDs and steroids for systemic use before the 12 weeks of gestatioin for VB-affected and VB-unaffected pregnancies and before the index date for terminations and miscarriages  ^c^ Analyses could not be adjusted for smoking in pregnancy since the data were unavailable for pregnancies ending in a termination or miscarriage  CI, Confidence interval; HR, Hazard ratio; VB, vaginal bleeding | | | | |

### **Table 4 Hazard ratios with 95% confidence intervals for any cancer and groups of site-specific cancers after additionally adjusting for BMI, Denmark, 2004-2018**

|  |  | **VB-affected vs VB-unaffected pregnancy^a^** |
| --- | --- | --- |
| **Outcome** | **Observations** | **HR (95% CI)** |
| Any cancer | All identifiable pregnancies | 0.96 (0.78-1.18) |
|  | First pregnancy | 1.23 (0.85-1.79) |
| Hormone-related cancers | All identifiable pregnancies | 0.62 (0.32-1.20) |
|  | First pregnancy | 0.94 (0.35-2.52) |
| Hematological cancers | All identifiable pregnancies | 1.13 (0.92-1.38) |
|  | First pregnancy | 0.84 (0.55-1.28) |
| Immune-related cancers | All identifiable pregnancies | 0.94 (0.47-1.90) |
|  | First pregnancy | 0.48 (0.07-3.46) |
| Smoking or alcohol-related cancers | All identifiable pregnancies | 0.77 (0.51-1.15) |
|  | First pregnancy | 0.50 (0.19-1.33) |
| Obesity-related cancers | All identifiable pregnancies | 1.07 (0.75-1.53) |
|  | First pregnancy | 1.19 (0.62-2.31) |
| Cancers of neurological origin | All identifiable pregnancies | 1.59 (0.85-2.99) |
|  | First pregnancy | 2.61 (1.06-6.41) |
| Other cancers | All identifiable pregnancies | 0.96 (0.78-1.18) |
|  | First pregnancy | 1.23 (0.85-1.79) |
| ^a^ Adjusted for age, calendar year of pregnancy end, women’s parity and number of previous identifiable pregnancies, socioeconomic factors (education, employment, and year-specific income level divided into quartiles based on all included pregnancies), smoking in pregnancy, BMI, reproductive history (pre-index date vaginal bleeding, termination, or miscarriages, placental complication in previous deliveries), history of chronic somatic conditions, any psychiatric condition, and use of NSAIDs and steroids for systemic use before the 12 weeks of gestatioin for VB-affected and VB-unaffected pregnancies and before the index date for terminations and miscarriages  BMI, body mass index; CI, Confidence interval; HR, Hazard ratio; VB, vaginal bleeding | | |

### **Table 5 Hazard ratios with 95% confidence intervals for breast, cervical, ovary and fallopian tube cancers after additionally adjusting for smoking in pregnancy, Denmark, 2004-2018**

|  |  | **VB-affected vs VB-unaffected pregnancy^a,b^** |
| --- | --- | --- |
| **Outcome** | **Model** | **HR (95% CI)** |
| Breast cancer | Crude | 0.95 (0.77-1.18) |
|  | Adjusted | 0.95 (0.77-1.18) |
| Cervix uteri cancer | Crude | 1.16 (0.82-1.65) |
|  | Adjusted | 1.13 (0.80-1.61) |
| Ovary and fallopian tube | Crude | 1.29 (0.53-3.13) |
|  | Adjusted | 1.19 (0.49-2.91) |
| Uteri corpus and unspecified | Crude | NA (NA-NA)^c^ |
|  | Adjusted | NA (NA-NA) ^c^ |
| ^a^ Computed using all identifiable pregnancies of a woman  ^b^ Adjusted for age, calendar year of pregnancy end, women’s parity and number of previous identifiable pregnancies, socioeconomic factors (education, employment, and year-specific income level divided into quartiles based on all included pregnancies), smoking in pregnancy, BMI, reproductive history (pre-index date vaginal bleeding, termination, or miscarriages, placental complication in previous deliveries), chronic somatic and psychiatric conditions, medication use before the index date as a proxy for overall underlying health and healthcare-seeking behaviour  ^c^ Non-available due to low number of accrued events  BMI, body mass index; CI, Confidence interval; HR, Hazard ratio; VB, vaginal bleeding | | |

### **Table 6 Hazard ratios with 95% confidence intervals for any cancer following the last identifiable pregnancy at age 40+ years, Denmark, 1995-2018**

| **Outcome** | **VB-affected vs VB-unaffected pregnancy^a^** | **VB-affected pregnancy vs termination^b^** | **VB-affected pregnancy vs miscarriage^b^** |
| --- | --- | --- | --- |
| **HR (95% CI)** | | | |
| **Any cancer** | 0.99 (0.79-1.25) | 0.94 (0.75-1.19) | 0.89 (0.70-1.12) |
| ^a^ Adjusted for age, calendar year of pregnancy end, women’s parity and number of previous identifiable pregnancies, socioeconomic factors (education, employment, and year-specific income level divided into quartiles based on all included pregnancies), smoking in pregnancy, reproductive history (pre-index date vaginal bleeding, termination, or miscarriages, placental complication in previous deliveries), history of chronic somatic conditions, any psychiatric condition, and use of NSAIDs and steroids for systemic use before the 12 weeks of gestatioin for VB-affected and VB-unaffected pregnancies and before the index date for terminations and miscarriages  ^b^ Analyses could not be adjusted for smoking in pregnancy since the data were unavailable for pregnancies ending in a termination or miscarriage  CI, Confidence interval; HR, Hazard ratio; VB, vaginal bleeding | | | |
